# Genotype-specific effects of physical activities on self-harm behavior in depressed patients: findings from the UK Biobank

**DOI:** 10.1101/2024.05.10.24307087

**Authors:** Jaegyun Jung, Sangyeon Lee, Jeong Ho Lee, Doheon Lee

**Author notes:** Corresponding author: Doheon Lee, PhD Affiliation: Department of Bio and Brain Engineering, Korea Advanced Institute of Science and Technology (KAIST), Daejeon, Republic of Korea Postal/mail address: #505, Chung Moon-Soul Building (E16), KAIST, 291 Daehak-ro, Yuseong-gu, Daejeon, Republic of Korea E-mail address, (Direct).

## Abstract

**Background:** Physical activities are widely implemented for non-pharmacological intervention in depression. However, there is little evidence supporting their genotype-specific effectiveness in reducing the risk of intentional self-harm in midlife and late-life depression patients.

**Aims:** To assess whether the specific type and amount of physical activity is associated with the risk of intentional self-harm behavior among patients with depression phenotype.

**Method:** We developed the bidirectional analytic model to investigate the genotype-specific effectiveness (BAIGE) on UK Biobank. After the genetic stratification on the depression phenotype cohort using hierarchical clustering, multivariable logistic regression models and Cox proportional hazard models were built to investigate the associations between physical activity and the risk of intentional self-harm behavior.

**Results:** A total of 28,923 subjects with depression phenotype were included in the study. In retrospective cohort analysis, the moderate and highly active groups were at lower risk of hospitalizations (moderate: adjusted odds ratio [OR], 0.77 [95% CI, 0.61-0.98], high: OR, 0.76 [95% CI, 0.60-0.97]). In the follow-up prospective cohort analysis, light-intensity physical activity (LIPA) was associated with a lower risk of hospitalizations in one genetic cluster (adjusted hazard ratio [HR], 0.26 [95% CI, 0.08 – 0.88]), which was distinguished by three genetic variants: rs1432639, rs4543289, and rs11209948. Compliance with the guideline-level moderate-to-vigorous physical activities was not significantly related to the risk of hospitalizations.

**Conclusions:** Physical activities, particularly a certain amount of light-intensity physical activity, mitigate the risk of hospitalizations due to self-harm in depression patients with a specific genotype.

## Introduction

Depressive disorder is one of the most prevalent mental disorders, affecting over 300 million people.^1^ The prevalence of depressive disorder is positively related to age, which makes middle-aged and older adults a population group at higher risk.^2^ Midlife and late-life depression are not only associated with cognitive impairment and dementia but also with intentional self-harm (ISH) behavior, including suicidal attempts. It is a widely held view that depression is the most common ‘antecedent’ of ISH. It is notable that self-harm is less prevalent in older adults, but when they commit ISH, it more often led to suicidal attempts. Older people commit suicide proportionally more than any other age group, and most of them suffer from depression.^3,4^ In addition, ISH is a common reason for hospitalizations. As suicide prevention is being a priority in public health strategy in many countries, there have been several attempts to investigate the impacts of hospitalizations due to intentional self-harm (HISH). According to the previous studies, HISH cost the total of 162 million GBP in the UK in 2017 and direct cost of 60 million USD for Denmark, and it is even increasing. Furthermore, these costs for inpatient care accounts for a large portion of these expenses.^5,6^ Taken together, the public health impact and use of medical resource highlights the need for study on HISH and its prevention.

Although the mechanism of ISH in patients with depression is still unclear, it has commonly been assumed that gene-environment interactions (GxEs) play a vital role in developing the condition.^5^ In consideration of this, non-pharmacological intervention, which might affect the environmental factors has begun to be widely used, along with the concern about the effectiveness of antidepressants.^6,7^

Physical activity, which refers to activity that involves bodily movement produced by skeletal muscles that requires energy expenditure, is one of the most commonly implemented treatment options in depression.^8,9^ A considerable amount of literature has reported the effectiveness of physical activity in reducing depressive symptoms, including suicidal ideation, while it varies by genotype.^10-12^

Physical activity may be classified into four main categories according to the absolute intensity, or rate of energy expenditure: sedentary, light, moderate, and vigorous activity.^13^ Despite its wide clinical use, there are still no clear recommendations on the level and intensity of physical activity tailored for depression patients to prevent serious consequences, such as HISH. For example, up-to-date guidelines from the World Health Organization (WHO) mostly focused on moderate-to-vigorous physical activity (MVPA) for the general population with little considerations on emerging and under-studied concepts such as light intensity physical activity (LIPA) or genotype-specific effect of physical activity.^9^

To address these evidence gaps, we examined the beneficial effect of physical activity on the risk of HISH in probable depression phenotypes. By utilizing the longitudinal nature and comprehensive data ranging from sociodemographic information to electronic health records (EHR) and accelerometry data, we sought to determine the optimal range and intensity of physical activity for midlife to late-life depression patients.

## Method

### UK Biobank

The UK Biobank project is a large prospective cohort data comprising 502,492 participants from England, Scotland, and Wales. Since 2006, extensive data including sociodemographic variables, online questionnaires, lifestyle factors, physical activity data, and genetic data, were collected on the participants who visited 22 assessment centers across the United Kingdom. The participants were aged 40-69 in recruitment, and the follow-up data until 2021 was included in this study. All the participants provided written informed consent, and the project has approval from the North West Multi-centre Research Ethics Committee (MREC) as a Research Tissue Bank (RTB) approval (REC reference: 21/NW/0157, IRAS project ID: 299116). Our research was conducted under application number 86585.

### Phenotype and outcome

A comprehensive medical record of each participant, coded clinical events, such as consultations, diagnoses, procedures, and laboratory tests, and registration records, including admission and discharge information, are linked to the UK Biobank project. In this study, primary healthcare data (Category 3000), summary diagnoses (category 2002), and record-level patient data (Category 2006) were used to define the depression cohort and outcome.

The depressive patients cohort was constructed according to a definition provided by Smith et al.: participants who include any of the following ICD-10 diagnoses - F32 (depressive episode), F33 (recurrent depressive episode), F34 (persistent mood [affective] disorders), F38 (other mood [affective] disorders), and F39 (unspecified mood [affective] disorder); or participants who were depressed/down for a whole week (Field ID 4598) for at least two weeks (Field ID 4609) and ever seen a GP or psychiatrist for nerves, anxiety or depression (Field ID 2090 and 2100); or participants with anhedonia for a whole week (Field ID 4631) which lasted for at least two weeks (Field ID 5375) and ever seen a GP or psychiatrist for nerves, anxiety, or depression (Field ID 2090 and 2100).^14^ Primary outcome of the study was ISH, which was defined using the hospital inpatient records of X60-X84 (intentional self-harm) diagnoses.

### Genotyping and imputation

Genome-wide genotype data was collected on all participants using either the UK Biobank Lung Exome Variant Evaluation Axiom or the UK Biobank Axiom array and underwent quality-control procedures performed by Wellcome Trust Centre for Human Genetics, University of Oxford, UK.^15^ Genotype data was imputed based on Haplotype Reference Consortium (HRC) and UK10K haplotype resource. Imputation data of over 90 million variants were available through the UK Biobank under approval. We selected 55 risk SNPs of major depressive disorder and depressive symptoms from published sources, which were available in imputed genotype data from the UK Biobank.^16-18^ Participants with any missing value in imputation data or mismatch in reported genetic and reported sex were excluded from the analysis.

### Physical activity

Two types of physical activity assessment were used in this study: subjective and objective measurement for the phase 1 and 2, respectively. For phase 1, we used Metabolic Equivalent Task (MET) score based on the latest version of International Physical Activity Questionnaire – Short Form (IPAQ-SF).^19^ After having adjusted for activity type (walking, moderate, and vigorous)-specific weight, MET score for each subject was derived and used to classify populations into three categories. Data was processed and categorized following the guideline published by IPAQ Research Committee.^20^

For objective assessment of the physical activity, E-mail invitations were sent to UK Biobank participants with valid e-mail addresses between February 2013 and December 2015. Over 100,000 participants have responded and agreed to provide the data and were sent Axivity AX3 wrist-worn triaxial accelerometer from May 2013, which was designed by Open Lab, Newcastle University. The participants were instructed to wear the device continuously for a whole week to track their entire physical activity.^21^

From measured raw acceleration data at 100Hz with a dynamic range ±8*g*, physical activity data were extracted after calibration procedures. We also classified the raw records of accelerometry data into four types of physical activity – sleep, sedentary behavior, LIPA, and MVPA – following the published methods of machine-learning-based behavior classification.^22^

### Statistical analyses

This study follows an ambidirectional cohort study design, utilizing the prospective and retrospective nature of the UK Biobank cohort. Sixteen years of longitudinal follow-up time period was divided into two phases: from 2006 to May 2013 for Phase 1 and from June 2013 to 2021 for Phase 2.

Genotype data were binarized, corresponding to the presence of the risk SNPs. Subjects are then clustered based on the Euclidean distance of the binarized genetic vector by hierarchical clustering. Since the number of clusters is unknown, we found the optimal threshold of hierarchical clustering using a silhouette coefficient. We generated decision trees to investigate significant SNPs for each cluster.

Univariate and multiple logistic regression analyses were conducted for the retrospective cohort study in phase 1 (from 2006 to May 2013). Assessing the relationship between the intentional self-harm behavior and self-reported physical activity level measured with IPAQ in each genetic cluster, the multivariable logistic regression model was adjusted for the potential confounders, including age group, sex, ethnicity, current employment status, education level, Townsend deprivation index, smoking status, and alcohol consumption.

The prospective cohort for phase 2 (from June 2013 to 2021) consists of participants without HISH medical records until the beginning of the phase. To assess the effectiveness of physical activity as a non-pharmacological intervention for depressive symptoms, the optimal level of each physical activity had to be decided prior to analysis. Previous guidelines by the World Health Organization (WHO) have indicated the recommended level of MVPA, whereas there is no such recommendation for LIPA.^9^ Therefore, the recommended level of LIPA was calculated using the methodology suggested by Chen et al.^23^ Then penalized cox-proportional hazard regression analysis was conducted in order to examine the associations between the types and guideline or calculation-based levels of physical activity and incident HISH in each genetic cluster.

We used Python 3.10.9 and SciPy 1.10.1 for genetic stratification, and ‘survival’ 3.5.5, ‘rms’ 6.7-1, and ‘CutpointsOEHR’ 0.1.2 with a R version 4.2.2 (R Foundation for Statistical Computing, Vienna, Austria) for other analyses. Mean and standard deviation (SD) are reported for continuous baseline characteristics except for the Townsend index (of which statistics are reported with median and interquartile range), and percentages are reported to present categorical variables. Odds ratios and hazard ratios are presented with a 95% confidence interval.

## Results

Among 64,826 participants who met the criteria for the probable depression phenotype, 36,238 were eligible for the imputed genotype-based stratification. After excluding the subjects with missing data in covariates, 28,923 depressive participants were included in the retrospective cohort study (eFigure 1 in Supplement). Participants without records of HISH until May 2013 and provided valid accelerometer data comprised the prospective cohort in Phase 2. Sociodemographic details and descriptive statistics of the cohort are presented in Table 1.

**Table 1.**
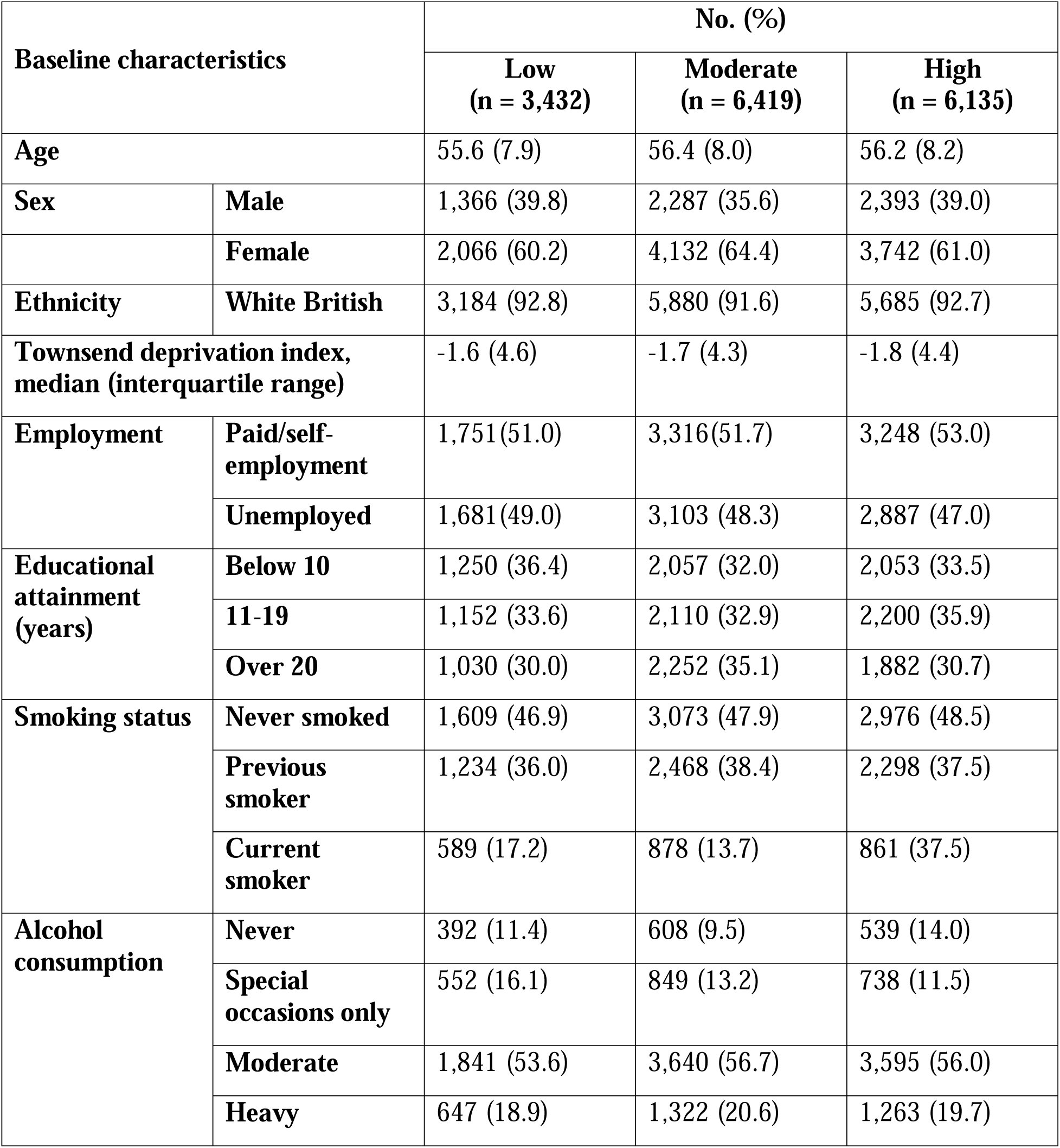
Sample characteristics of the participants according to the IPAQ physical activity group.

### Genetic stratification

We found the three largest clusters from the result of hierarchical clustering; each consists of 6,681, 8,905, and 4,401 participants (Figure 1). Also, genetic features that distinguish each cluster were found through the decision tree algorithm. Following cohort studies were conducted targeting these three clusters.

**Figure 1.**
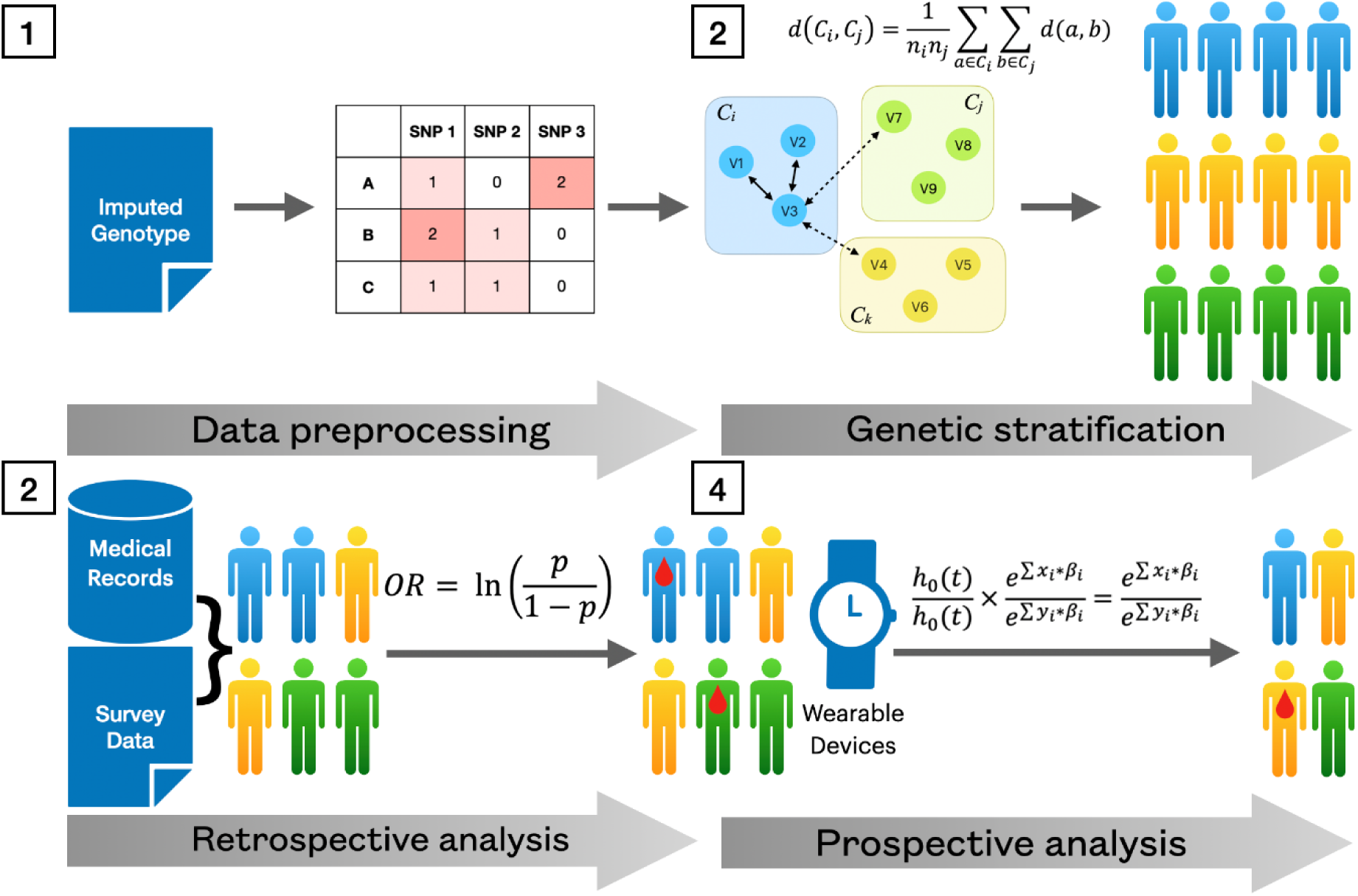
Scheme of the bidirectional analytic model to investigate the genotype-specific effectiveness (BAIGE)

### Phase 1: Retrospective cohort study

In phase 1, we sought to examine the association between the IPAQ physical activity group and HISH risk in each genetic cluster. Overall, in unadjusted logistic regression models, participants in moderate or IPAQ physical activity groups were associated with significantly lower odds of HISH. There were similar findings in the models stratified with summed MET minutes per week for all activity. Multivariable models which were adjusted for covariates showed that more active IPAQ groups were at the lower risk odds of HISH in the full cohort (OR for the moderately active group: 0.771[0.609-0.977], OR for the highly active group: 0.764[0.602-0.971]), although there were mixed findings from each genetic cluster (Table 2).

**Table 2.** Unadjusted and adjusted odds ratios (ORs) for risk of HISH according to the IPAQ physical activity group in the three largest genetic clusters.

| **Genetic cluster** | **N** | **IPAQ group** | **Model 1**<br>**OR (95% CI)** | **Model 2**<br>**aOR (95% CI)<sup>a</sup>** |
| --- | --- | --- | --- | --- |
| 1 | 5,365 | Low | Reference |  |
|  |  | Moderate | 0.60<br>(0.41 – 0.88) | 0.71<br>(0.47 – 1.05) |
|  |  | High | 0.65<br>(0.45 – 0.96) | 0.77<br>(0.52 – 1.15) |
| 2 | 7,086 | Low | Reference |  |
|  |  | Moderate | 0.57<br>(0.39 – 0.82) | 0.70<br>(0.48 – 1.01) |
|  |  | High | 0.70<br>(0.49 – 1.00) | 0.85<br>(0.59 – 1.22) |
| 3 | 3,535 | Low | Reference |  |
|  |  | Moderate | 0.84<br>(0.53 – 1.36) | 1.03<br>(0.64 – 1.69) |
|  |  | High | 0.49<br>(0.29 – 0.83) | 0.61<br>(0.35 – 1.04) |
| Total | 15,986 | Low | Reference |  |
|  |  | Moderate | 0.64<br>(0.52 – 0.80) | 0.77<br>(0.61 – 0.98) |
|  |  | High | 0.64<br>(0.50 – 0.80) | 0.76<br>(0.60 – 0.97) |

a. The aORs from Model 2 are adjusted for all covariates listed in Table 1.

### Phase 2: Prospective cohort study

Beginning in June 2013, we assessed the relationships between the accelerometer-measured physical activity levels and incident HISH in 7,137 participants during a mean (SD) follow-up period of 8.55 (0.44) years. After the classification of the accelerometer data, the cohort was stratified using the guideline-based thresholds for MVPA (≥150 minutes per week). Since the amount of LIPA had a U-shaped relationship with the log-relative hazards, the ‘goldilocks’ level for LIPA (between 2.38 and 9.53 hours a day) was derived by calculated optimal cut points (Figure 2).

**Figure 2.**
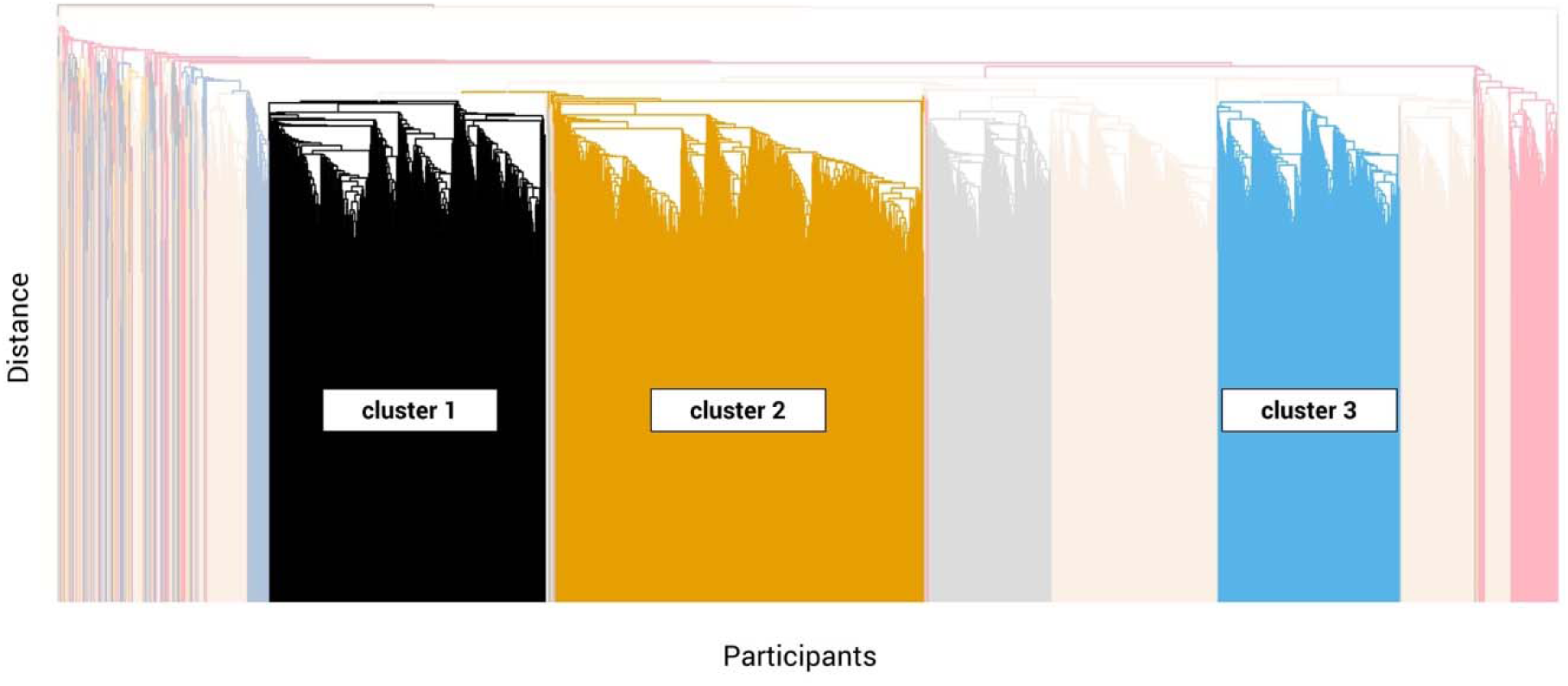
Dendrogram for the hierarchical agglomerative clustering.

**Figure 3.**
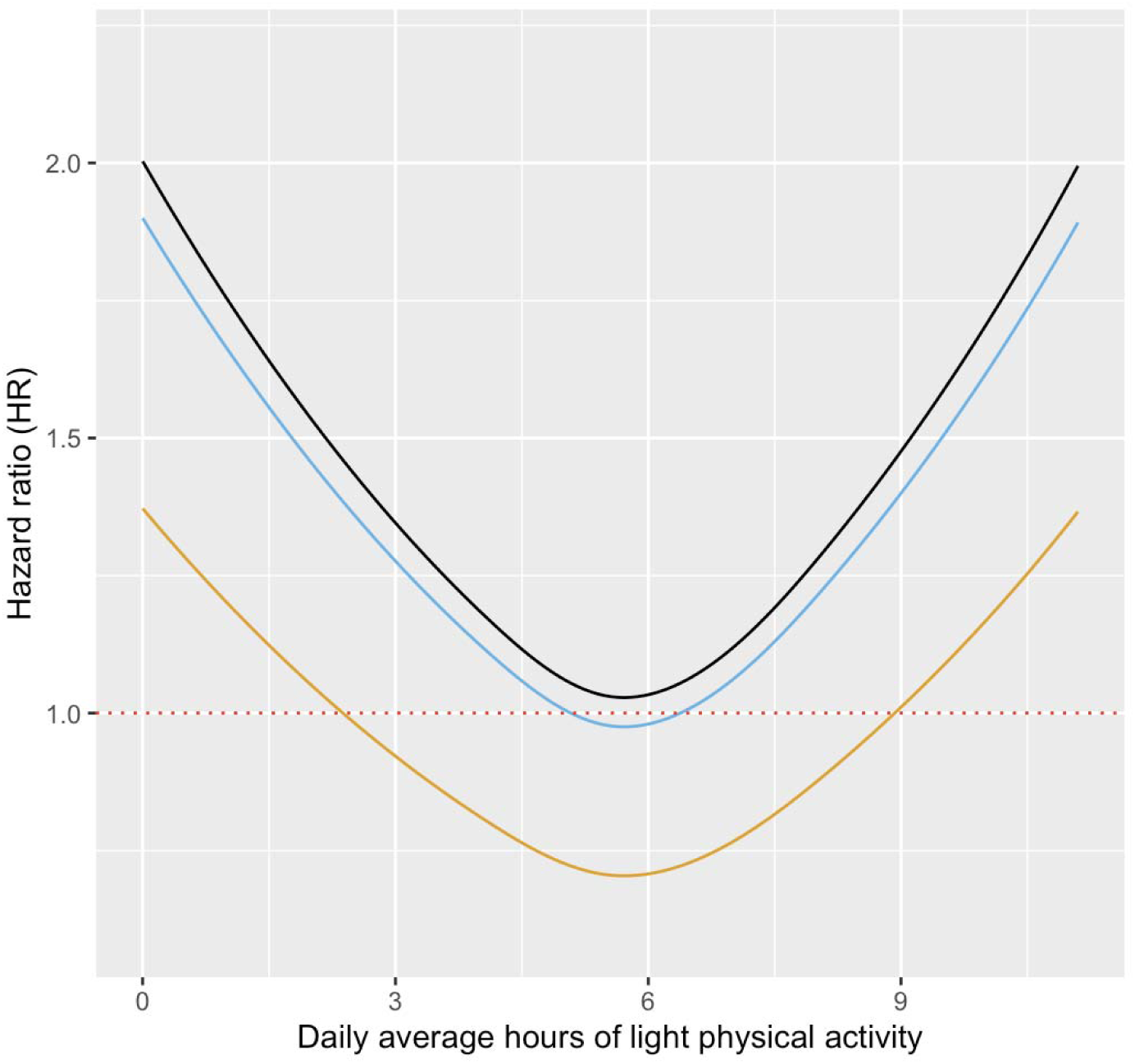
The ‘U-shaped’ relationship between the daily average hours of light-intensity physical activity and hazard ratios (HRs) of HISH.

The results of penalized cox-proportional hazard regression analysis are shown in Table 3. ‘Goldilocks’ amount of LIPA was associated with a lower risk of HISH in genetic cluster 2 (HR, 0.23; 95% CI, 0.07 – 0.76), and it was significant after adjusting for possible confounders (adjusted HR, 0.26; 95% CI, 0.08 – 0.88), although there were no significant differences observed in other clusters. No significant differences in risk of HISH were found between the levels of MVPA.

**Table 3.** Unadjusted and adjusted hazard ratios (HRs) for HISH in group within recommended level of light-intensity physical activity (LIPA) and moderate-to-vigorous physical activity (MVPA)

| **Genetic Cluster** | **No. of events (%)** | **No. of participants** | **Model 1 HR (95% CI)** | **Model 2 adjusted HR (95% CI)** |
| --- | --- | --- | --- | --- |
| **Light-intensity Physical Activity<sup>a</sup>** |  |  |  |  |
| 1 | 13 (1.1) | 1,204 | 0.61 (0.14 – 2.69) | 0.65 (0.15 - 2.93) |
| 2 | 11 (0.7) | 1,563 | 0.26 (0.08 – 0.84) | 0.26 (0.08 - 0.88) |
| **Moderate-to-vigorous Physical Activity (MVPA)** |  |  |  |  |
| 1 | 10 (1.3) | 746 | 1.53 (0.52 – 4.49) | 1.18 (0.39 – 3.59) |
| 2 | 9 (0.9) | 983 | 1.42 (0.48 – 4.24) | 1.38 (0.44 – 4.30) |
| 3 | 7 (1.3) | 528 | 1.14 (0.33 – 3.89) | 0.85 (0.24 – 2.97) |

a. The results for cluster 3 was not applicable due to the insufficient number of events. No significant difference was found between the group with or within LIPA recommendation in the log-rank test ( $p = 0.3$ ).
b. The adjusted HRs from Model 2 are adjusted for all covariates listed in Table 1.

## Discussion

The current study sought to examine the preventive effect of physical activity on the HISH of a population with depression phenotype. The findings from the retrospective cohort study in phase 1 revealed that depressive participants who engage in an average or higher level of physical activity tend to be at lower risk of HISH. In the subgroup analysis, the relationship between the overall physical activity level and incident HISH differed across the genetic clusters. A possible explanation for the mixed results and discrepancies across the genetic clusters is the genotype-specific effect of physical activity, which in turn heightens the need for detailed research on the detailed classification of physical activity and its impact in groups with different genotypes.

Analysis of the prospective cohort settings in phase 2 showed that participation in machine-learning derived ‘goldilocks’ level of LIPA reduces the risk of HISH in genetic cluster 2. Since the effect was not significant in other clusters, it can thus be suggested that the calculated level of LIPA could be recommended for patients with specific genetic variants for cluster 2.

Meanwhile, the data from this study did not confirm the significant effect of guideline-based MVPA thresholds in the prevention of HISH. These findings are broadly consistent with recent studies that found the beneficial effect of LIPA on depressive symptoms rather than MVPA. Although existing studies and recommendations have focused on MVPA or the overall amount of physical activity, recent reports have suggested that objectively measured LIPA is more effective than another type of physical activity in alleviating depressive symptoms.^24-27^ Because of their limitations in compliance and adherence to the physical activity prescription, LIPA could be a more appropriate option for the middle-aged and older population.^25^

Although the molecular mechanism of LIPA’s preventive effect on depressive symptoms is not clear, one can assume that LIPA affects the pathophysiology of depression in the way general physical activity does. According to our analyses, there were three SNPs that distinguish cluster 2: rs1432639, rs4543289, and rs11209948. Previous results from GWAS reported that two of the three variants, rs1432639 and rs11209948, are related to obesity/BMI and body fat percentage, respectively.^18,28^ The nearest gene of the two variants is neuronal growth regulator 1 (NEGR1), which is suggested to be a central hub that links depression and obesity.^29^ When considering the proposed mechanism of neuroplasticity of physical activity in depression, the two variants possibly involved in mediating neural plasticity along with NEGR1.^30^ Taken together with the role of rs4543289 in severity and symptoms of depression, the complex interaction between these genetic components could provide a probable explanation for the greater effect of physical activity on HISH in cluster 2.^17^

One of the distinguishing features of our research is that we utilized various study designs and measurement tools available to extend the current findings. For example, throughout our study, the amount of physical activity was largely associated with a relatively lower risk of incidence HISH. Although the association was inconsistent in the retrospective cohort study, which made use of categorization derived from self-reported questionnaires, we took advantage of objectively measured physical activity and confirmed effectiveness in the prospective cohort settings. Recent evidence, ranging from the Mendelian randomization study and meta-analyses, has reported the possible discrepancies between subjective and objective measurement of physical activity, and objective measurement (i.e., accelerometer data) is more likely to be related to the improved outcomes in depression and related outcomes.^31-33^ This discrepancy could be attributed to the bias of self-reported assessment tools. In spite of its convenience and cost-effectiveness, subjective assessment of physical activity is vulnerable to measurement error and various types of bias. These limitations could be more consequential when it comes to recall and social desirability bias in respondents in older age, or with mental disorders.^24,34^ Moreover, it has been reported that participation in LIPA is especially prone to recall bias, which in turn provides a possible explanation for the lack of previous research on it and emphasizes the need for epidemiological studies with objective physical activity assessment.^35^

To the best of our knowledge, this is the first study to examine the genotype-specific effect of LIPA on HISH in midlife and late-life depression. While depression in midlife and late-life is becoming more prevalent, serious consequences of the disorder, such as ISH, is recognized as a major public health concern. Furthermore, research on personalized intervention for depression has been mostly restricted to pharmacological treatment.^36^ The present findings have implications for developing personalized recommendations for LIPA, while further research on more genotypes and psychophysiological factors is needed for a better understanding of the non-pharmacological treatment options for depression.

### Limitations

There are several limitations to this study. First, this study was based on a single database, which in turn could be affected by possible biases such as ethnicity in the original dataset. Second, most of the variables were collected at a specific time point, which means that the variables might not fully represent the characteristics of the participants. For example, participants might have changed their physical activity pattern or lifestyle factors during the observation period or after the measurement. Third, the genetic stratification was based on previously reported risk SNPs, so there might have been other genetic variants that we have yet to account for, and would have uniquely found in the UK Biobank dataset. Fourth, since the sum of hours during a day is fixed, the effects of specific intensity and level of activity might partly be due to change in other types of activities. For example, previous studies and guidelines reported the benefit of displacing the sedentary activity to LIPA or MVPA. Fifth, because the statistical model used in the analysis presupposes a ‘U-shaped’ relationship between the continuous variable and the log-relevant hazard ratio, the optimal range could not be calculated for MVPA, of which the relationship with the hazard ratio was not ‘U-shaped.’ However, the findings of this study are consistent with previous reports when applying guideline-based thresholds.^24^

## Data Availability

The data supporting the findings of this study are available from UK Biobank (https://www.ukbiobank.ac.uk/) and can be requested under license.

