## Supplementary material for "Genotype-specific effects of physical activities on self-harm behavior in depressed patients: findings from the UK Biobank"

**Supplementary Materials**

**Supplementary Fig 1.** Study Flow

**Supplementary Table 1.** Definition of the Covariates

**eReference**

Supplementary Fig 1. Study Flow

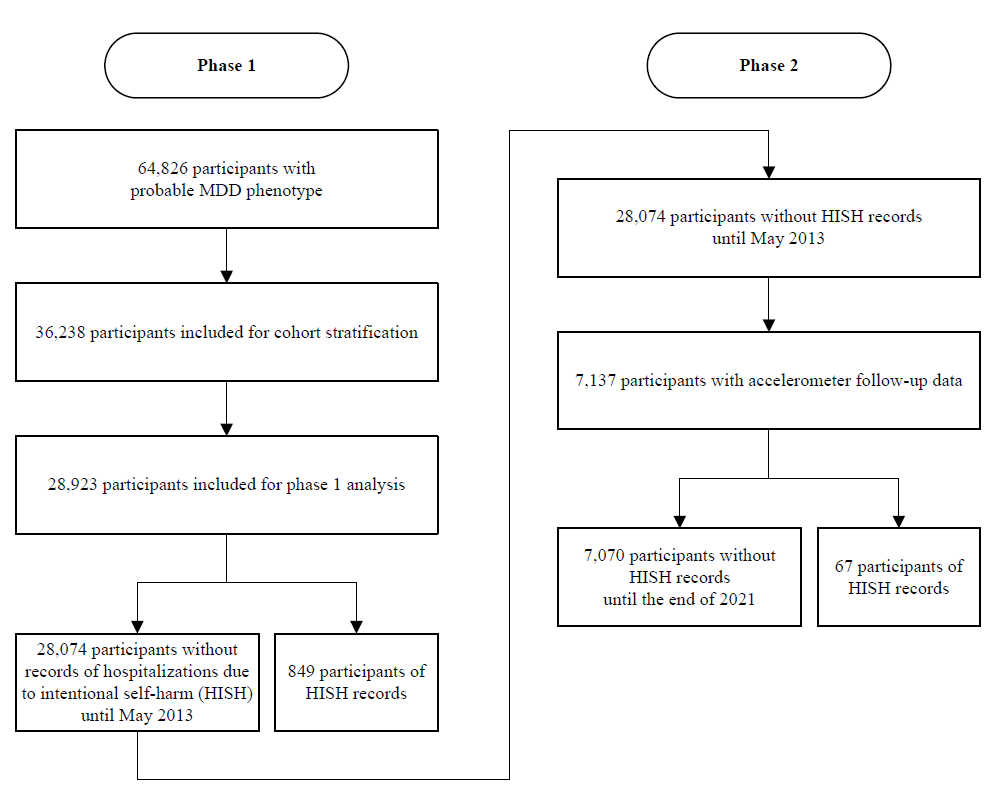

Supplementary Table 1. Definitions of the covariates

| **Covariates** | **UK Biobank Data-Field ID** | **UK Biobank Data-Field Description** | **Coding** | **Notes** |
| --- | --- | --- | --- | --- |
| Age group | 21022 | Age at recruitment | Continuous age in years | Coded in three categories: 40-50, 50-60, >60 |
| Sex | 31 | Sex | 1) Female  2) Male |  |
| Ethnicity | 21000 | Ethnic background | 1) White  2) Mixed  3) Asian or Asian BritHISH  4) Black or Black BritHISH  5) Chinese  6) Other ethnic group | Dichotomized into White British and others |
| Townsend deprivation index | 22189 | Townsend deprivation index at recruitment | Continuous value |  |
| Employment status | 6142 | Current employment status (Instance 0) | 1) In paid employment or self-employed  2) Retired  3) Looking after home and/or family  4) Unable to work because of sickness or disability  5) Unemployed  6) Doing unpaid or voluntary work  7) Full or part-time student | Coded in two categories: ‘Employed (In paid employment or self-employed)’ and ‘Not employed (other answers)’ |
| Education level | 6138 | Qualifications (Instance 0) | 1) College or University degree  2) A levels/AS levels or equivalent  3) O levels/GCSEs or equivalent  4) CSEs or equivalent  5) NVQ or HND or HNC or equivalent  6) Other professional qualifications eg: nursing, teaching  7) None of the above | Years of education was derived from the highest qualification that each subject achieved, according to the International Standard Classification for Education (ISCED) definitions.(1) Education level was classified as high (more than 20 years of education), intermediate (11 to 19 years of education), and low (less than 10 years of education). |
| Smoking status | 20116 | Smoking status (Instance 0) | 1) Never  2) Previous  3) Current | Dichotomized into 1) Never smoked and 2) Ever smoked |
| Alcohol consumption | 1558 | Alcohol intake frequency (Instance 0) | 1) Daily or almost daily  2) Three or four times a week 3) Once or twice a week  4) One to three times a month  5) Special occasions only  6) Never | Classified into four categories: 1) Never, 2) Special occasions only, 3) Moderate (one time a month to four times a week), and 4) Heavy (Daily or almost daily) |
